# Treatment and Eradication of *Helicobacter Pylori* in a Safety-Net Health System

**DOI:** 10.1101/2025.09.25.25336669

**Authors:** Rena Mei, Erin Libuit, Rachel Corren, Omar Viramontes, Dalia Martinez, Shreya Patel, Justin L. Sewell, Ma Somsouk

## Abstract

**Background & Aims:** *Helicobacter pylori* (HP) is the most common chronic bacterial infection worldwide, with a rising resistance rate, although the resistance patterns among minority and immigrant populations are not known. We aimed to determine the failure rate by treatment and race/ethnicity among adult patients with HP infection in a safety-net health system.

**Methods:** We conducted a retrospective cohort study in a safety-net health system from January 2020 to December 2021 of patients diagnosed with HP based on serology, stool antigen, or stomach biopsies who were prescribed antibiotic therapy and had a test of eradication by either stool antigen or histology. The primary outcome was the HP treatment failure rate according to treatment type. Additional subgroup analyses examined the failure rate by age, race/ethnicity, and preferred language.

**Results:** 167 patients carried a diagnosis of HP that was treated with a test of eradication. The median age was 47 (range 18-79); 60% were female, 52% Hispanic, and 25% Asian. 93% received a 14-day course, and 84% were prescribed clarithromycin-based triple therapy. The overall *H. pylori* treatment failure rate was 18%. The failure rate for quadruple therapy was nominally higher compared to triple therapy (29.2% vs. 16.3%, p=0.13). Younger patients (age ≤ 45) tended to experience treatment failure (24% vs. 13%, p=0.07). Asian (7.3%, p=0.01) and Cantonese-speaking patients (4.2%, p=0.02) had the lowest overall failure rates.

**Conclusions:** In this study of diverse adult patients, triple therapy tended to be more effective than quadruple therapy. Triple therapy was particularly effective for Asian patients. These findings raise the possibility that triple therapy could still serve as a first-line regimen in certain populations.

## Introduction

*Helicobacter pylori* (*H. pylori*, or HP) is the most common chronic bacterial infection worldwide, with more than half of the world’s population infected^1^. It is a Gram-negative bacterium that colonizes the human stomach, where it causes chronic inflammation. It disrupts the gastric mucous layer and attaches itself to the gastric epithelium, leading to altered gastric secretion and tissue injury from the host immune response. Although many people remain asymptomatic, over time this can lead to chronic gastritis, peptic ulcer disease, and gastric malignancies, including carcinoma and lymphoma.

Treatment of HP includes a combination of antibiotics and acid-suppressing medications. However, the antibiotic resistance rates of HP have been rising^2,3^, and studies have demonstrated that pre-treatment HP antibiotic resistance is a major factor that markedly increases treatment failure^4,5,6^. As such, the updated 2024 American College of Gastroenterology (ACG) clinical guidelines recommend bismuth quadruple therapy as first-line therapy in treatment-naïve patients over clarithromycin-based triple therapy^7^. The aim of our study was to examine prescribing patterns in an integrated safety-net system that serves a diverse population and to compare the real-world failure rate of quadruple therapy compared to triple therapy, including across race/ethnicity and other clinical subgroups.

## Methods

### Study setting and population

A retrospective cohort study was performed in the San Francisco Health Network (SFHN), an integrated safety-net health system in San Francisco, California. Approval was obtained from the Institutional Review Board at the University of California, San Francisco (IRB #20-30255). The San Francisco Health Network (SFHN) is a community of clinics, hospitals, and programs operated by the San Francisco Department of Public Health, providing healthcare to all San Franciscans regardless of immigration status or insurance coverage, essentially acting as the city’s public health system serving diverse populations. Many patients are first generation immigrants from Asia and Latin America. We included adult patients above 18 years of age with a diagnosis of HP infection by serology, stool, or histology who were prescribed treatment with antibiotic therapy, and who had confirmed eradication testing by either stool antigen or histology between January 2020 and December 2021. Antibiotic therapy regimens were divided into triple therapy or quadruple therapy. Patients were excluded if they were diagnosed with HP but did not receive treatment, or received treatment but did not have confirmation testing.

### Analytic Plan

The primary outcome was the failure rate of treatment, with treatment failure defined by persistence of HP infection based on biopsy or stool antigen. Descriptive statistics and chi-squared tests were performed to compare the failure rate of triple therapy versus quadruple therapy. Additional sub-group analyses examined the failure rate by age, race/ethnicity, and preferred language. Treatment failure was further analyzed according to the method of HP diagnosis, grouped by serology versus stool antigen or histology.

## Results

A total of 167 patients were included in the analysis based on the inclusion and exclusion criteria (Table 1). Baseline characteristics of the patients included a median age of 47 (range 18-79) and 60% female. The majority of patients were Hispanic (52%), followed by Asian (25%); 9% were non-Hispanic white and 7% non-Hispanic Black. Only 28% of patients spoke English as their preferred language, compared to 47% Spanish and 19% East Asian languages. 85% of patients received triple therapy as first-line treatment compared to 15% who received quadruple therapy. 83.8% of treatment regimens were clarithromycin-based triple therapy and 13.2% were bismuth quadruple therapy. 93% received a 14-day antibiotic regimen compared to 7% who received a 10-day regimen.

**Table 1.**
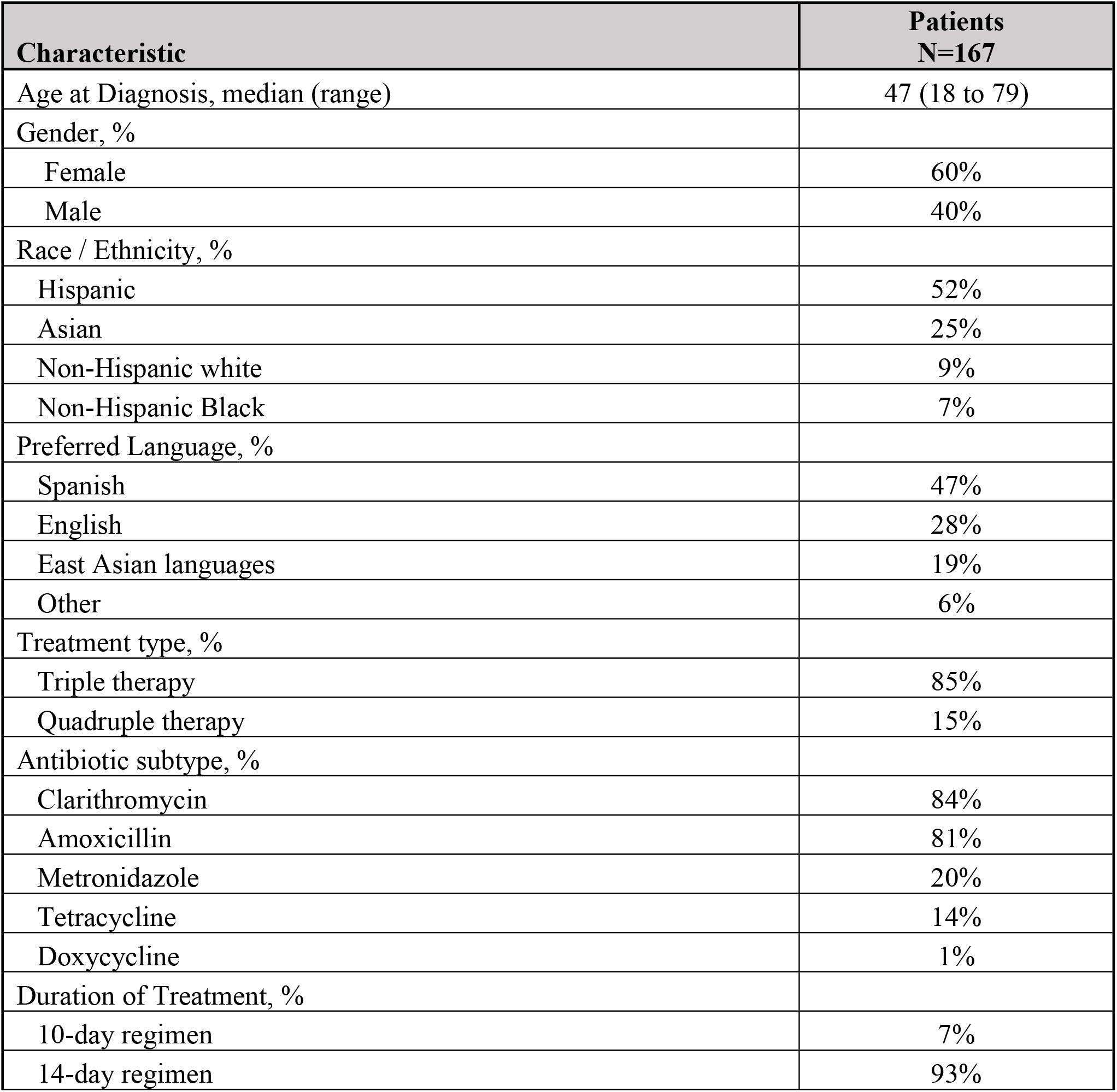
Baseline characteristics of patients with confirmed *H. pylori* infection.

The overall HP treatment failure rate was 18% (Table 2). The failure rate of quadruple therapy was 29.2% compared to 16.3% for triple therapy (p=0.13). The failure rate of clarithromycin was 15.6% compared to 30.4% failure rate of tetracycline (p=0.08). The failure rate for females was 20.8% compared to 13.6% in men (p=0.24) (Table 3). Younger patients (age ≤ 45) were somewhat more likely to experience treatment failure (24% vs. 13%, p=0.07). Asian patients (7.3%, p=0.01) and Cantonese-speaking patients (4.2%, p=0.02) had the lowest overall failure rates. There was no significant difference in failure rates of 10-day regimens compared to 14-day regimens (18.2% vs 17.9%, p=0.98).

**Table 2.** Rates of *H. pylori* treatment failure by treatment type.

| Category | Treated, n | Failed, n | Failure Rate (%) | P-value |
| --- | --- | --- | --- | --- |
| **Overall** | 167 | 30 | 18.0% |  |
| **Treatment type** |  |  |  |  |
| Triple therapy | 141 | 23 | 16.3% |  |
| Quadruple therapy | 24 | 7 | 29.2% | 0.13 |
| **Antibiotic subtype** |  |  |  |  |
| Clarithromycin | 141 | 22 | 15.6% |  |
| Amoxicillin | 135 | 21 | 15.6% | 0.99 |
| Metronidazole | 34 | 9 | 26.5% | 0.14 |
| Tetracycline | 23 | 7 | 30.4% | 0.08 |
| Doxycycline | 1 | 0 | 0.0% |  |
| **Duration of Treatment** |  |  |  |  |
| 14-day regimen | 156 | 28 | 17.9% |  |
| 10-day regimen | 11 | 2 | 18.2% | 0.98 |

**Table 3.** Rates of *H. pylori* treatment failure by patient characteristics.

| Category | Treated, n | Failed, n | Failure Rate (%) | P-value |
| --- | --- | --- | --- | --- |
| **Age** |  |  |  |  |
| < = 45 | 75 | 18 | 24.0% |  |
| > 45 | 92 | 12 | 13.0% | 0.07 |
| **Gender** |  |  |  |  |
| Female | 101 | 21 | 20.8% |  |
| Male | 66 | 9 | 13.6% | 0.24 |
| **Race / Ethnicity** |  |  |  |  |
| Hispanic | 87 | 23 | 26.4% |  |
| Asian | 41 | 3 | 7.3% | 0.01 |
| Native Hawaiian/Pacific Islander | 6 | 1 | 16.7% | 0.6 |
| Non-Hispanic black | 12 | 1 | 8.3% | 0.17 |
| Non-Hispanic white | 15 | 2 | 13.3% | 0.28 |
| **Preferred Language** |  |  |  |  |
| Spanish | 78 | 21 | 26.9% |  |
| English | 47 | 6 | 12.8% | 0.06 |
| Cantonese | 24 | 1 | 4.2% | 0.02 |
| Other | 18 | 2 | 11.1% | 0.16 |

The treatment failure rate tended to be higher in patients who had their HP diagnosed via stool antigen or stomach biopsy compared to serology (21.6% vs. 11.6%, p=0.09). When HP was diagnosed by a stool antigen or stomach biopsy, treatment failure remained generally lower for triple therapy compared to quadruple therapy (19% vs. 36%, p=0.19). Similarly, for patients who had an HP diagnosis by serology, treatment failure was nominally lower for triple therapy (14% vs. 20%, p=0.61).

## Discussion

In this retrospective cohort study of adult patients at a safety net hospital with confirmed HP infection, triple therapy tended to be more effective than quadruple therapy. Triple therapy was more commonly prescribed and appeared to be more effective, with an estimated failure rate of 16.3% compared to quadruple therapy with a failure rate of 29.2%. Asian patients and those whose preferred language was Cantonese experienced the lowest rate of treatment failure, at less than 10%, with the vast majority prescribed a triple therapy.

The effectiveness of triple therapy observed in this study urges caution with the suggestion by the 2024 ACG clinical guidelines recommending bismuth quadruple therapy as the first-line treatment for HP. The updated recommendation was made in response to rising clarithromycin resistance rates and studies demonstrating superiority of bismuth quadruple therapy over clarithromycin-based triple therapy within and outside of the United States^3,8,9^. Indeed, in a large cohort study from the United States and Europe, clarithromycin resistance was estimated at 22.2%^3^, while a meta-analysis suggested a pooled resistance rate of 31.5% in the United States^2^. However, pooled clarithromycin resistance rates varied among ethnic groups, with a rate of resistance of 17.1% for Black patients but nearly double the rate for white patients (29.9%), Hispanic patients (31.2%), and others (40.8%). There are also wide variations in resistance rates based on country of origin. A study in east Los Angeles found much higher resistance rates to clarithromycin based on susceptibility testing of gastric biopsies among patients from Mexico and Central/South America compared to those born in the United States (26% and 22% versus 9%)^10^. Despite estimates of high clarithromycin resistance in the population, our study suggests that either or both resistance rates and eradication rates for triple therapy are better compared to quadruple therapy. Indeed, a 2018 study in a diverse patient population in New Jersey also supported the use of clarithromycin-based triple therapy as a first-line treatment, with eradication rates of 84-86% within the practice^11^. This suggests that despite the rising rates of macrolide resistance at the population level, there may be other factors that impact treatment failure, such as adherence and varying exposure to macrolide antibiotic by geography. In our safety-net population, it is possible that clarithromycin resistance rates may not be high, or if it is high, HP may still be eradicated with adequate adherence. The Asian subgroup suggests that failure rates may be as low as about 5%. Thus, although ACG clinical guidelines favor bismuth quadruple therapy as first-line therapy for HP, one should consider the local rates of treatment failure. In addition, the choice of treatment among a diverse safety-net population should take into account the circumstances impacting patients’ ability to adhere to a more complex treatment course and their exposure to prior antibiotics.

There were several limitations to this study. A larger sample size is needed to account for confounding in triple therapy compared to quadruple therapy by gender and ethnicity. We included positive HP serology tests in the diagnosis of HP to increase the power of the study, but we acknowledge that this test cannot differentiate between active and cured infection, as observed by the lower rate of treatment failure when HP was diagnosed by serology. This may account for why the overall failure rate in this study (18%) was slightly lower than the published literature, which is described to be 19.8% in a real-world observational cohort analysis of insured patients in the U.S. between 2016 and 2019^12^. Additionally, the data did not include information on whether each patient was taking a proton pump inhibitor at the time of the test of eradication, which can affect the positivity rate as proton pump inhibitors can lead to false negative test results. Lastly, we excluded patients who did not have a test of eradication, which could bias the results towards higher rates of treatment failure since test of eradication is more likely pursued in patients with complications of HP.

In summary, triple therapy appears to be more effective than quadruple therapy in this real-world analysis in one safety net health system. Future efforts should increase the sample size to confirm this finding, assess variations by immigration status and geography, examine treatment failure patterns in other settings, and correlate treatment effectiveness with antibiotic resistance patterns.

## Data Availability

see below

## Abbreviations

(ACG): American College of Gastroenterology
(HP): *Helicobacter pylori*
(SFHN): San Francisco Health Network

